# Short-range exposure to airborne virus transmission and current guidelines

**DOI:** 10.1101/2021.04.06.21255017

**Authors:** Jietuo Wang, Mobin Alipour, Giovanni Soligo, Alessio Roccon, Marco De Paoli, Francesco Picano, Alfredo Soldati

## Abstract

After the Spanish flu pandemic, it was apparent that airborne transmission was crucial to spreading virus contagion, and research responded by producing several fundamental works like the experiments of Duguid [J. Hyg. 44:6, 1946] and the model of Wells [Am. J. Hyg., 20:611–18,1934]. These seminal works have been pillars to past and current guidelines published by health organizations. However, in about one century, understanding of turbulent aerosol transport by jets and plumes has enormously progressed and it is now time to use this body of developed knowledge. In this work, we use detailed experiments and accurate computationally-intensive numerical simulations of droplet-laden turbulent puffs emitted during sneezes in a wide range of environmental conditions. We consider the same emission – number of drops, drop size distribution and initial velocity – and we change environmental parameters as temperature and humidity, and we observe strong variation in droplets evaporation or condensation in accordance with their local temperature and humidity microenvironment. We assume that 3% of the initial droplet volume is made of non-volatile matter. Our systematic analysis confirms that droplets lifetime is always about one order of magnitude larger compared to previous predictions, in some cases up to 200 times. Finally, we have been able to produce original virus exposure maps, which can be a useful instrument for health scientists and practitioners to calibrate new guidelines to prevent short-range airborne disease transmission.

**Significance Statement:** Violent expiratory events represent an important route for the spread of respiratory viruses, as the SARS-CoV-2 virus. We use finely-resolved experiments and simulations to quantify how the turbulent cloud of moist air exhaled during a sneeze largely increases the airborne time and the lifespan of virus-loaded droplets. By providing visualizations of the spatial distribution of the virus copies, we highlight the high infection risk associated with droplets that remain airborne in the near proximity of an infected individual. The present study aims at raising awareness among public health authorities about this infection risk, which is grossly underestimated by current guidelines.

---

Respiratory viruses can be transmitted among human subjects via three main routes. First, direct contact and fomites, where a healthy individual comes into direct contact with an infected person (direct contact) or touches a contaminated surface (fomite). Second, through the droplet transmission that occurs in the proximity of an infected person, who exhales large and small respiratory droplets containing the virus. Third, through the airborne transmission of smaller droplets and particles (droplet nuclei), which remain airborne over a much longer time, traveling farther distances than droplet transmission. While the latest research suggests that direct contact and fomites are unlikely to be a major source of infection for SARS-CoV-2 (1, 2), understanding the role played by the latter two contributions is crucial to design effective guidelines for pathogens transmission prevention.

Conventionally, and according to WHO guidelines (3), droplets larger than 5 *µm* in diameter are referred to as respiratory droplets (droplet transmission), while those smaller than 5 *µm* in diameter are defined as droplets nuclei (airborne transmission). This threshold has been widely used to define public health guidelines and to design infection control interventions for healthcare workers (4). The current pandemic, however, highlighted the limitations of these guidelines and made it clear that new research should be embraced to revise these recommendations. Indeed, the threshold used to distinguish between droplet transmission and airborne transmission and its scientific rationale are highly questionable (5, 6): the aerodynamic behavior of droplets, ballistic for respiratory droplets and aerosol-like for droplet nuclei, strongly depends on the local flow conditions. Recent works suggest that a much larger threshold (100 *µm*)(7, 8) better differentiates between large and small droplets dynamics. Likewise, the better understanding of turbulence gained in the last 50 years, has shown how the evaporation process is extremely complex (9–11) and cannot be captured with simplified models, like those employed by Wells (12). Finally, from a medical perspective, it is evident how the distinction between large droplets and small droplets (airborne) diseases and its connection with the short- and long-range transmission is rather weak and for many respiratory infections the predominant route depends on the specific setting (13, 14).

A key step towards understanding the routes of pathogens transmission must rely on the study of fluid dynamics as it plays a crucial role in almost every aspect of disease spreading (10, 15). Thanks to recent experimental and numerical advancements, we can access to detailed time- and space-resolved quantities. In this work, using the most recent experimental and numerical methodologies, we investigate the evaporation and dispersion dynamics of respiratory droplets in four different ambient conditions (temperature and relative humidity). Building on these results, and using virological data, we evaluate the transport of the viral copies providing graphical visualizations of the infection risk at close distance from an infected subject. Our findings suggest that predictions based on the models adopted in current guidelines are largely unsatisfactory, leading to a dangerous underestimation of the infection risk. In particular, current guidelines underestimate the infectious potential associated with the short-range airborne route (6, 16, 17), i.e. the infection risk associated with small droplets and droplet nuclei that remain airborne in the proximity of an infected individual and that may readily penetrate and deposit in the upper and lower respiratory tract (18).

## Results and Discussion

We start by comparing the behavior predicted by our experiments and simulations of a sneezing event; these results are then benchmarked against theoretical scaling laws available for the two phases characterizing the expiratory event: jet and puff (19–21). Then, high-fidelity simulations are used to characterize the dispersion and evaporation of the respiratory droplets in different ambient conditions. These results are compared with the predictions obtained from models currently employed in public health guidelines. Finally, we try to bridge fluid dynamics and virological data on SARS-CoV-2 to characterize the virus exposure discussing the risk associated with droplets of different sizes.

### Sneezing event: simulations and experiments

To assess the reliability of numerical simulations in accurately reproducing a sneezing event, we start by benchmarking simulation results against those obtained from the experiments performed in the TU Wien laboratory. Due to the impossibility of performing experiments in which temperature, vapor mass fraction and velocity fields are recorded simultaneously, we focus on the ability of simulations and experiments to accurately capture the flow structures and the dynamics of the sneeze. To this aim, we consider a neutrally buoyant jet having the same temperature and humidity as the ambient (*T* = 22 °C and *RH* = 50%) seeded with nearly mono-disperse droplets having a diameter equal to 2 *µm* (tracers). For simulations, the inflow condition is obtained from a gamma-probability-distribution function (22), which mimics the airflow generated by a sneezing event. Likewise, experiments have been designed to reproduce an inlet condition that is repeatable and similar to that adopted in the simulations. Please refer to *Materials and Methods* and *SI Appendix* for further details.

To quantitatively compare the results, we consider the time evolution of the jet front. Results are also benchmarked against the theoretical scaling laws available in the literature (19–21). In particular, considering the finite duration of a sneezing event (and consequently the finite time during which momentum is injected in the environment), we can distinguish between two different phases: *i*) jet phase, linked to the early jet evolution when momentum is continuously provided (constant momentum flux); *ii*) puff phase, linked to the late evolution where momentum injection ceases and the jet momentum remains constant. Using the self-similarity hypothesis, two scaling laws for the distance traveled by the jet front can be derived (23); for the starting jet phase (constant momentum flux), the distance traveled by the jet front *L* grows over time as *L ∝ t*^1*/*2^, while for the puff phase (constant momentum), the penetration distance grows as *L ∝ t*^1*/*4^.

Figure 1 shows the evolution of the front of the jet obtained from simulations (red points) and experiments (blue points with error bars). As reference, the theoretical scaling laws for the jet and puff phase are reported with two dashed lines. For the jet phase, we observe a very good agreement between simulations and experiments, and the respective scaling law. The very small discrepancy observed between our results and the theoretical scaling law can be traced back to the constant momentum flux hypothesis used to derive the scaling. This assumption is only partially satisfied as in the very first stage of the sneeze (*t <* 0.1 *s*), there is a rapid, but not instantaneous, increase of the inlet velocity and thus of the momentum flux. Moving to the puff phase, there is a remarkable decrease of the momentum flux and the distance traveled by the jet front deviates from the jet scaling law and approaches the puff scaling law. Even in this later stage, an overall good agreement is observed among experiments, simulations and the theoretical scaling law. To further compare simulations and experiments, we measured the semi-cone angle of the jet obtaining very similar values among experiments (*α* = 8.5°) and simulations (*α* = 8°). These findings are also in agreement with previous investigations on human respiratory activities (20). Additional comparisons between simulations and experiments are available in the *SI Appendix*.

**Fig. 1.**
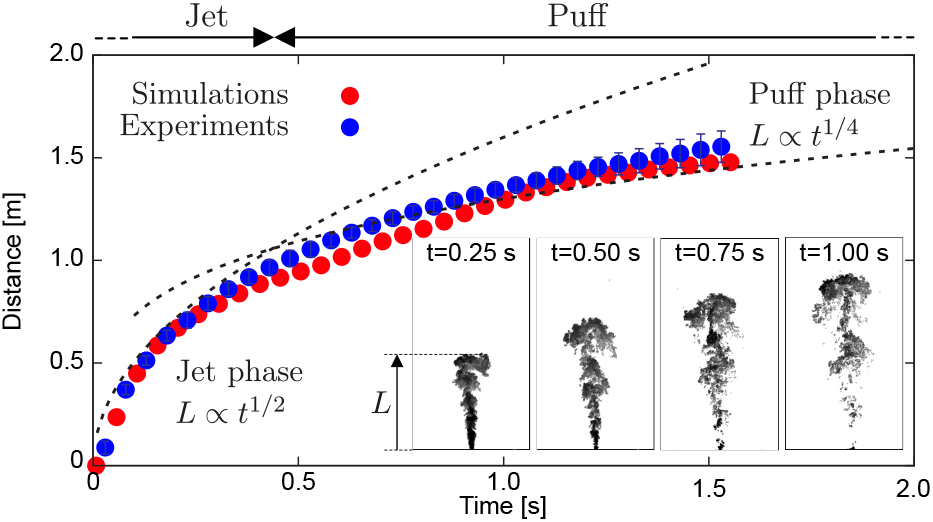
Distance traveled by the front of the jet: comparison between simulations (red dots) and experiments (blue dots). For experiments, data are obtained from 7 independent realizations and error bars corresponding to the standard deviation are also shown. The two stages that characterize the sneezing event, jet (early stage) and puff (late stage) are clearly visible; the scaling laws for the jet, *L ∝ t*^1*/*2^, and puff phase, *L ∝ t*^1*/*4^, are reported as reference with black dashed lines. Both simulations and experiments exhibit a very similar behavior and are in excellent agreement. Qualitative visualizations obtained from experiments showing the instantaneous tracers concentration (black-high; white-low) at different times (*t* = 0.25 *s, t* = 0.50 *s, t* = 0.75 *s* and *t* = 1.00 *s*, respectively) are reported as representative of the jet/puff evolution.

### Sneezing event simulations

Once assessed the reliability of the numerical framework, we use numerical simulations to study the dispersion and evaporation of respiratory droplets resulting from a sneeze. We study four different environmental conditions: two temperatures (*T* = 5 °C and *T* = 20 °C) and two relative humidities (*RH* = 50% and *RH* = 90%). These simulations are performed using the same numerical setup discussed before (see *Materials and Methods* and *SI Appendix* for details).

Figure 2 shows a graphical representation of the sneezing event reproduced by the simulations; the left column (A and B) refers to the *T* = 5 °C and *RH* = 90% case while the right column (C, D) refers to the *T* = 20 °C and *RH* = 50%. For each case, two different time instants, *t* = 0.25 *s* and *t* = 0.50 *s*, are shown; the background is colored by the local *RH* (white-low; black-high), while the dimension (not in scale) and color of the respiratory droplets correspond to their diameter (red-small; white-large). At the beginning (A, C), for both cases, most of the droplets are within the turbulent saturated cloud emitted by the sneezing jet. Only few droplets, with diameter larger than 100 *µm* (white), located in the front of the jet leave the cloud. Later in time (B, D), the largest droplets start to settle down and thus to significantly move along the vertical direction. On the contrary, most of the other droplets remain suspended in the vapor cloud generated by the sneezing jet as their settling time is longer (e.g. 600 *s* for a 10 *µm* droplet)(12). The effects of buoyancy is also apparent. Since the jet is characterized by a higher temperature (smaller density) than the environment, the cloud starts to move upwards carrying small droplets as well. This effect is more evident for the low temperature cases. Finally, it is worth to observe that already after 0.5 *s* the front of the jet with the transported droplets has already travelled about 1 *m* away from the infected individual.

**Fig. 2.**
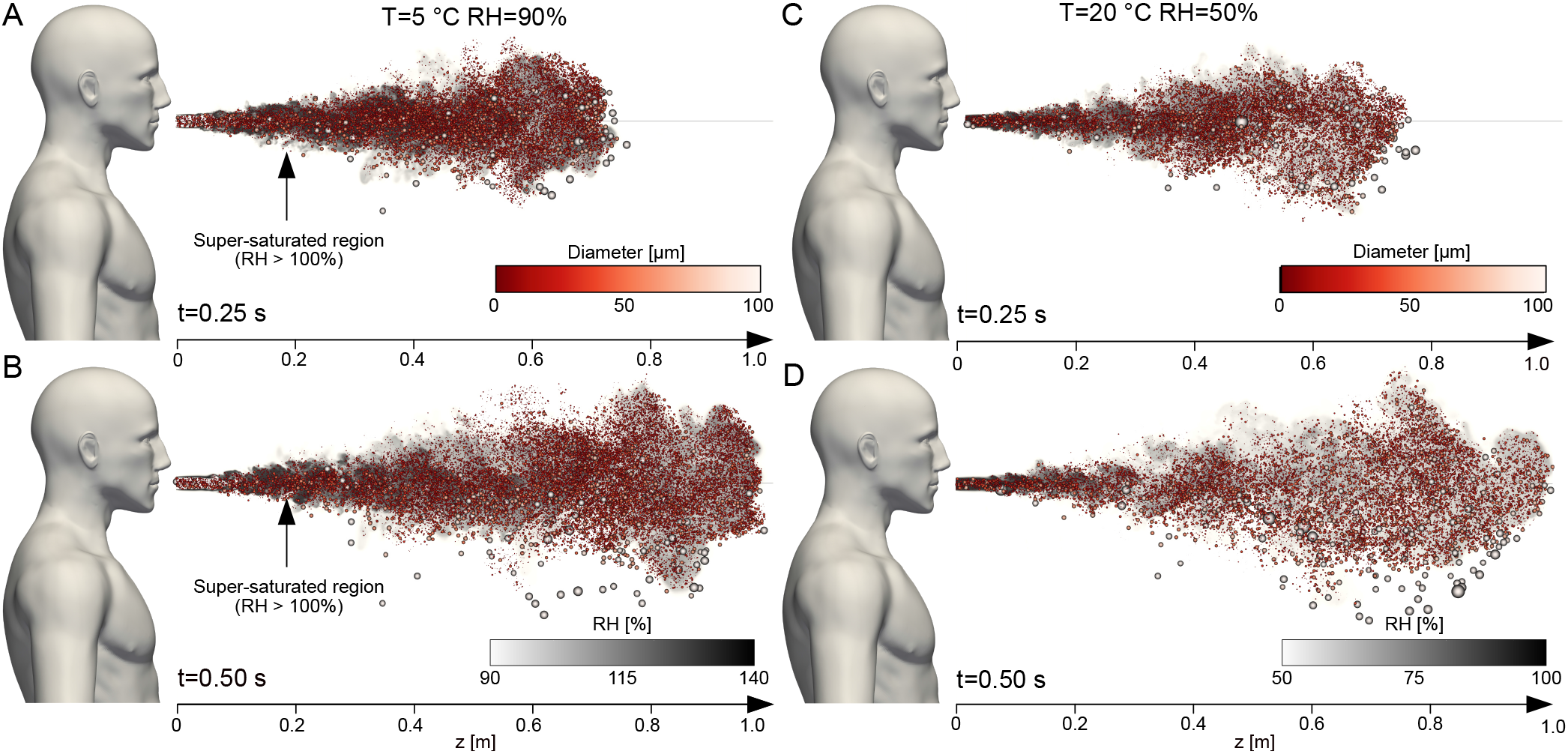
Snapshots of the sneezing event at: *t* = 0.25 *s* (A, C) and *t* = 0.50 *s* (B, D), where *t* = 0 represents the beginning of the respiratory event. The left column refers to *T* = 5 °*C* and *RH* = 90%, while the right column to *T* = 20 °*C* and *RH* = 50%. The background shows the local value of the relative humidity (white-low; black-high). The respiratory droplets are displayed rescaled according to their size (not in real scale) and are also colored according to their size (red-small; white-large). We can appreciate how most droplets move together with the turbulent gas cloud generated by the sneezing jet. This cloud is characterized by a much larger value of the RH with respect to the ambient. In addition, for *T* = 5 °*C* and *RH* = 90% (left column), a wide region is characterized by supersaturated conditions (*RH >* 100%).

### Evaporation of respiratory droplets

To evaluate the infection risk associated with droplets of different sizes, we first evaluate the lifetime of the respiratory droplets. To this aim, we compute the time required by each droplet to complete the evaporation process, reaching its terminal size determined by the presence of non-volatile elements. Indeed, since respiratory liquid contains salt and proteins (24–27), droplets evaporate until they reach a critical size forming droplet nuclei (water and non-volatile evaporation residua), which may remain suspended. The volume fraction of non-volatile elements varies between individuals and is on average about 3% in volume (24–28), which correspond to a dry nuclei size of about 30% of the initial droplet diameter. Figure 3 shows the resulting evaporation times for the different ambient conditions tested (A-D). The evaporation times are reported according to the initial droplet diameter and, for each class of diameters, we compute the probability of obtaining a given evaporation time. The sample plot on the left side of the figure guides the reading of the other four panels. For any given initial diameter, the leftmost part of the distribution marks the shortest evaporation time, while the rightmost part of the distribution marks the longest evaporation time; empty black circles identify the mean evaporation time for each initial diameter. The distribution is colored by the probability of each evaporation time (blue-low probability; yellow-high probability). Present results have been compared with the evaporation time predicted by the constant temperature model (12, 29), which is currently employed in most public health guidelines. This model, assuming an isolated droplet at constant ambient temperature, leads to the so-called *d*^2^-law, which predicts that the evaporation time is proportional to the initial diameter squared (and thus to the initial droplet surface). The predicted evaporation time (or more precisely the time required for a droplet to shrink down to 30% of its initial diameter) is reported with a red solid red line as a function of the diameter: according to the *d*^2^-law, small droplets evaporate almost immediately, while a much longer time is required for larger droplets. We highlight how the evaporation process obtained from simulations is different from that predicted by the model. Indeed, according to the *d*^2^-law, only droplet nuclei should be present beyond the red line, which marks the evaporation time predicted with the constant temperature model. However, simulations show a complete different picture with most of the droplets completing the evaporation process well beyond the predicted time. The slower evaporation dynamics of respiratory droplets is very pronounced for the high relative humidity cases (B,D), where only droplets smaller than 20 *µm* fully evaporate within 2.5 *s*. It is worth mentioning that for the *T* = 5 °C and *RH* = 90% case, the presence of a supersaturated region induced by the warm humid exhaled air (see figure 2A-B), produces an initial condensation of smaller droplets. Hence, in the first phase, droplets grow in size (11) instead of evaporating and shrinking.

**Fig. 3.**
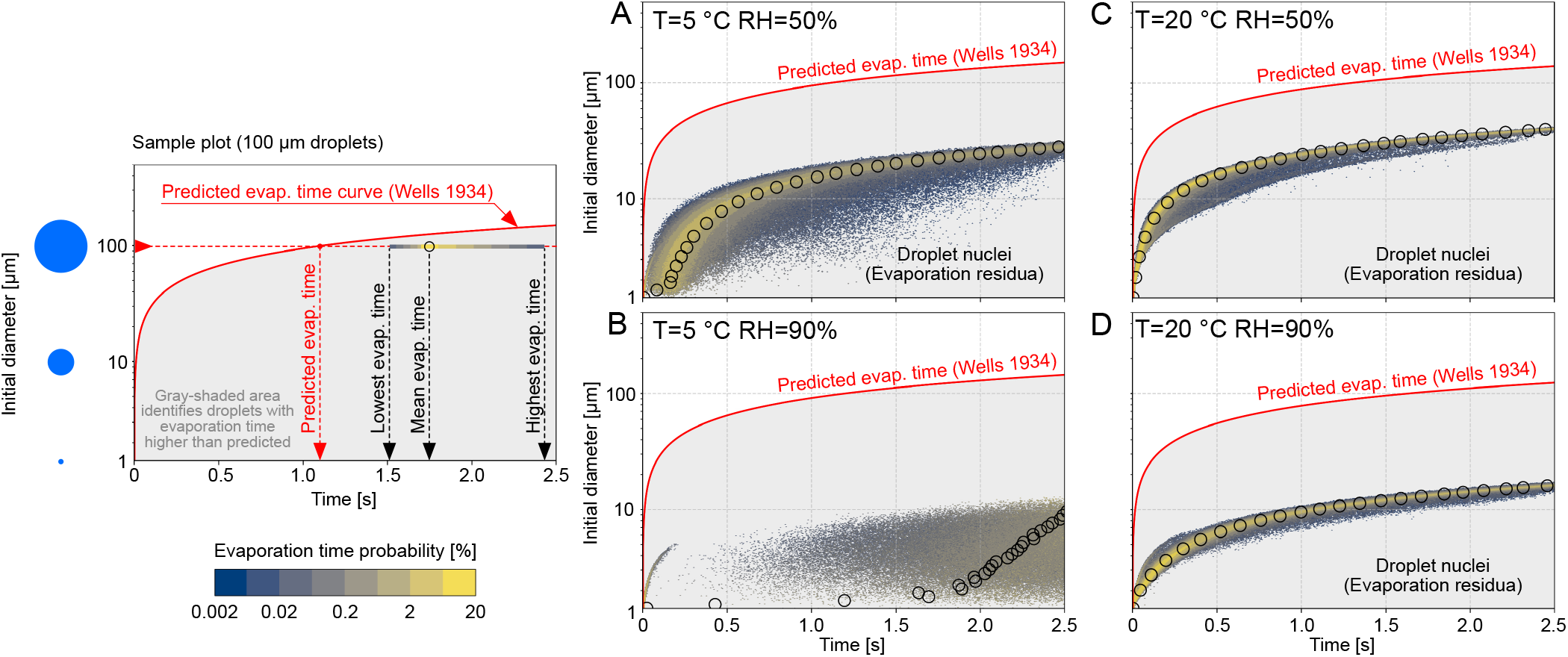
Time required by the respiratory droplets to complete the evaporation process in the four ambient conditions tested: *T* = 5 °C and *RH* = 50% − 90% (A and B) and *T* = 20 °C and *RH* = 50% − 90% (C, D). The sample plot on the left provides at a glance guidance on how to read the main panels. In particular, for any given initial diameter, the leftmost side of the distribution indicates the shortest evaporation time, while the rightmost side of the distribution marks the longest evaporation time observed for droplets with a certain initial diameter. The color of the distribution represents the probability (blue-low; yellow-high) of having a certain evaporation time. Empty black dots represent the mean evaporation time obtained from the simulation data. The predicted evaporation time obtained from the *d*^2^-law, a model currently employed for the definition of public health guidelines, is reported with a solid red line as a function of the droplet diameter. According to the model prediction, all droplets should evaporate within the time prescribed by the red line, thus the gray-shaded area below the red line should be empty (i.e. droplets should have already evaporated to dry nuclei). Simulations results, however, show a complete different picture and for all ambient conditions, droplets evaporate well beyond the predicted time. This reflects the action of turbulence and of moist air released during the sneeze, which largely slows down the evaporation. These effects are very pronounced for the low temperature/high humidity case (B), where only a fraction of droplets smaller than 10 *µm* completely evaporate to a dry nucleus within 2.5*s*. For the other cases (A, C and D), small droplets (less than 20 to 40 *µm*) complete the evaporation process, and the formation of droplet-nuclei can be appreciated.

A similar trend, but less marked, can be observed for the low humidity cases, where for the most favorable case (C), almost all droplets with a size smaller than 40 *µm* reach their final size. The resulting mean evaporation times obtained from simulations are thus larger (by at least one order of magnitude) than those predicted from the constant temperature model, as also observed in recent studies (11). This dramatic slowdown of the evaporation process traces back to the motion of the droplets and to the local thermodynamic conditions they sample. Indeed, most of the droplets are exposed to the warmer and more humid conditions that characterize the exhaled cloud (19, 30) and to their fluctuations produced by turbulence (21). As the evaporation rate of the droplets is determined by the local humidity value at the droplet position, which is much higher than the expected environmental value, this results in a much slower evaporation (i.e. a much longer evaporation time).

Besides, the formation of the droplet-nuclei can be also appreciated. For any given diameter, evaporated droplets shrink down to the critical size determined by the presence of non-volatile matter (30% of the initial diameter). The dispersion of these nuclei is critical in disease transmission as they carry a relatively large amount of bacteria and viruses (14, 31), which may remain infectious for a considerable amount of time, traveling long distances (e.g. for SARS-CoV-2, the half-life in aerosol is 1 ≃ *h*). The presence of droplet nuclei is observed in those cases characterized by a faster evaporation dynamics (A, C and D), while for the case *T* = 5 °C and *RH* = 90% (D), nuclei formation is strongly delayed as the evaporation process is hindered by the higher local relative humidity.

### Virus exposure maps

To evaluate the infection risk, we present the virus exposure maps. Specifically, we compute the cumulative number of virus copies that go past a control area in different domain locations. This type of evaluation requires precise information on the viral load for the SARS-CoV-2 virus. However, in archival literature, this information is characterized by large uncertainty and the reported viral loads differ by several orders of magnitude (32–38). Indeed, viral load measurements are not only influenced by the method used to test the swab, but viral load exhibits also strong variations during the different stages of the infection (36, 37, 39), being influenced by the severity of symptoms (40–42) and many other factors as well, among which age, sex and droplet size (37, 43, 44). To bypass this uncertainty, we present our results in a dimensionless form, normalized by the total number of virus copies ejected and assuming a uniform viral load across all droplets at the time of their ejection. The dimensional concentration of virions can be calculated by multiplying the data from the normalized virus exposure maps for the viral load (an indicative value is 7 × 10^6^ *copies/mL* for an individual with severe symptoms)(35, 36) and the ejected liquid volume (≃ 0.01 *mL*). As the time scale of present simulations (3 *s*) is much smaller than the half-time life (45, 46) in aerosol of SARS-CoV-2 (about 1 *h*), we do not consider any viral load decay.

Figure 4 shows the virus exposure obtained from the four ambient conditions tested (A-D). For all simulated cases, we observe a core-region (green) characterized by a rather uniform value of exposure and some hotspots characterized by larger values of exposure, up to ten times more. This behavior is attributed to the contemporary presence of droplets of very different sizes. Large droplets (more than 100 *µm*) carry a high number of virus copies (proportional to the initial droplet volume) and produce hotspots as their number is low; by opposite, small droplets (less than 100 *µm*) carry a lower number of virus copies and produce a more uniform exposure level as their number is higher. The presence of exposure hotspots (and thus of droplets larger than 100 *µm*) extends up to 1.25 *m*. Indeed, these larger droplets follow almost ballistic trajectories and soon settle to the ground. The core region is surrounded by an outer region characterized by a smaller level of exposure, that extends farther in space. This outer region is generated by smaller droplets and droplet nuclei, which reach this outer region later in time (*t >* 1 *s*) when the majority of the larger droplets have already settled to the ground and most of the smallest ones have completed the evaporation process (see figure 3).

**Fig. 4.**
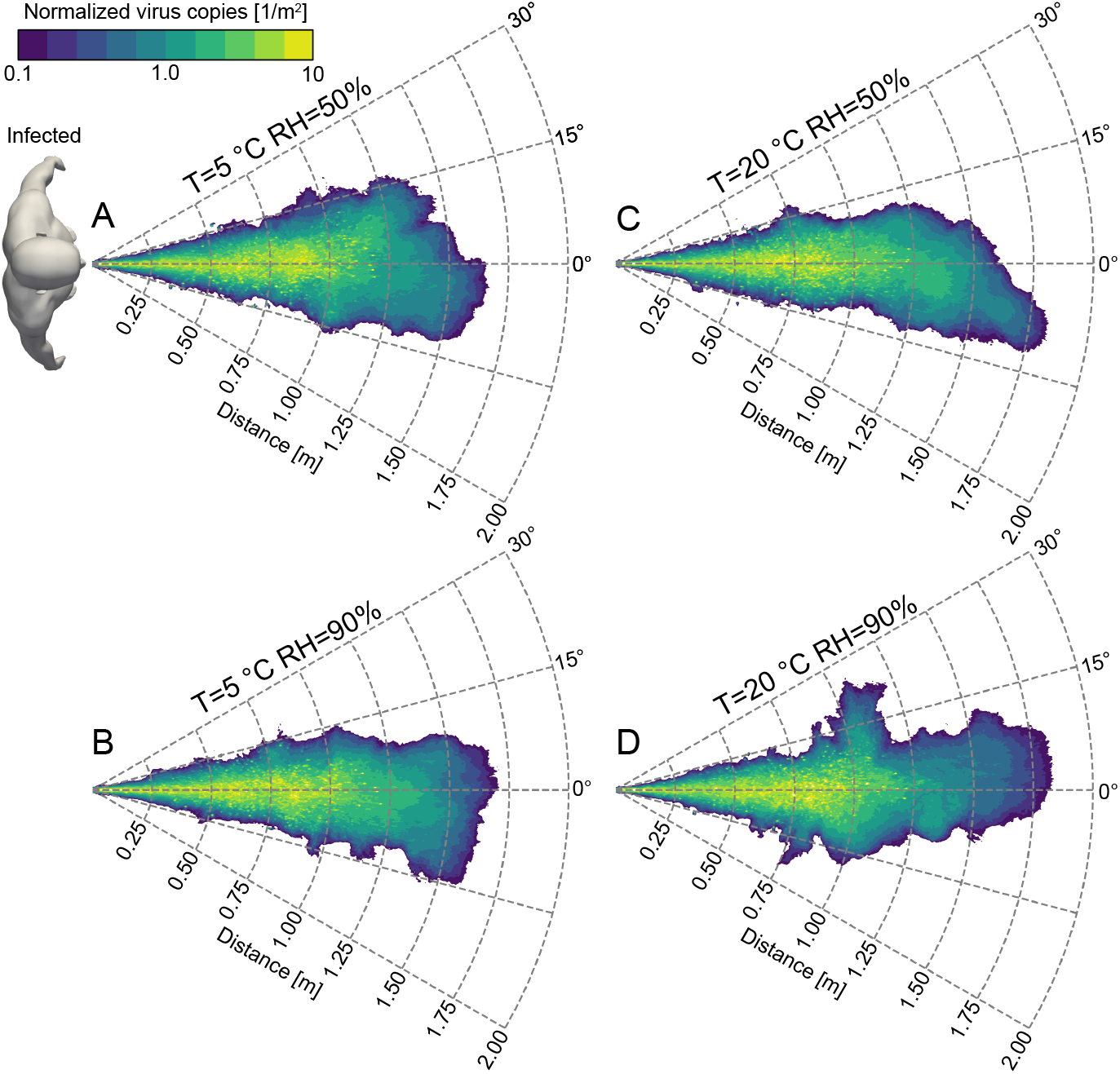
Virus exposure (violet-low; green-high) for the four ambient conditions simulated: *T* = 5 °C and *RH* = 50% − 90% (A-B) and *T* = 20 °C and *RH* = 50% − 90% (C-D). Exposure is defined as the number of virus copies (virions) that go past a control area in different locations of the domain. The results are shown normalized by the total number of virus copies ejected during a sneeze. The dimensional concentration of virus copies can be obtained by multiplying the normalized exposure data for the viral load and the ejected liquid volume (≃0.01 *mL* in the present simulations). We can observe the presence of a core region charac-terized by a high level of virus exposure, which is mainly determined by the large droplets (100 microns or more). These droplets follow almost ballistic paths and settle to the ground within ≃ 1.25 *m*. This core region is surrounded by a wider region characterized by a lower level of virus exposure. Although in this outer region the value of exposure is smaller, a susceptible individual is still exposed to thousands of virus copies (here we consider an average viral load for SARS-CoV-2 of 7 × 10^6^ *copies/mL*). According to the independent action hypothesis, the presence of thousands of virus copies in the small droplets and droplet nuclei poses a significant threat on both the short- and long-range airborne transmission routes of SARS-CoV-2.

Overall, although for smaller droplets, the probability of containing a virus copy is lower due to their initial small volume, we can observe how their large population leads to a remarkable level of virus exposure in the core region (hundreds of thousands of virions per square meter for a viral load of 7 × 10^6^ *copies/mL*) as well as in the outer region (thousands of virions per square meter for the same viral load). As the independent action hypothesis, which states that each pathogen individual has a non-zero probability of causing host infection, seems to apply for SARS-CoV-2, the virus exposure produced by these small droplets poses a significant risk for airborne transmission. It is interesting to observe that the risk of infection via small droplets (i.e. via the airborne route) is significant in the long-range (beyond 1 *m* from the source), but is even more important in the short-range where a remarkable level of exposure can be addressed to small droplets. This observation suggests that the airborne route has an important, if not dominant, role also in the short-range transmission, and it is not limited to the long-range route, as commonly assumed in most of the current guidelines. The potential of these small droplets in causing infections in the long-range is of difficult estimation as it depends on the virus viability in the droplet nuclei (45) and to the specific environmental conditions (e.g. wind, ventilation). However, considering the relatively high density of virus copies present in these small droplets and droplet nuclei, the risk of infection via the airborne route also in the long-range cannot be neglected. This risk is particularly pronounced in closed places where air dilution is low, as also documented by the large number of outbreaks that occurred in closed spaces (47–50).

## Conclusions

In this study, with the help of finely time- and space-resolved experiments and simulations, we provide evidence that current guidelines, which rely on recommendations based on seminal works (12, 51), present several flaws. A first flaw is identified in the standard prediction of the evaporation times: models currently used in public health guidelines grossly underestimate by at least one order of magnitude the actual evaporation times. A second flaw is represented by the threshold used to differentiate between large and small droplets; while this definition can be meaningful in healthcare environments where the term aerosol refers to a specific group of operations, from a fluid dynamics perspective, this criterium is questionable as the behavior of droplets is influenced by the local flow conditions (e.g. breath/cough/sneeze) and even large droplets (60*/*100 *µm*, considered as ballistic in most guidelines) stay suspended in the environment for a considerable amount time. These flaws lead to a consistent underestimation of the infection risk: formation of droplet-nuclei is delayed with respect to predictions and droplets remain in the most infectious condition (liquid) for a much longer time. In addition, the flow conditions generated by violent expiratory events (sneeze), allow 60*/*100 *µm* droplets to remain airborne for a significant amount of time. As highlighted by the virus exposure maps, this leads to a remarkable risk of infection via airborne particles also in the short-range transmission. In light of the present findings, we believe that mitigation of the infection risk via the short-range airborne route (6, 16, 17) must be addressed in current guidelines.

## Data availability

The datasets analyzed during the present study are available from the corresponding author upon reasonable request.

## Materials and Methods

We summarize here the numerical and experimental methodologies used. Further details on the numerical method, experimental setup and additional tests can be found in *SI Appendix*.

### Simulations

The numerical simulations are based on an hybrid Eulerian-Lagrangian framework. An Eulerian Large-Eddy-Simulation approach is used to describe the velocity, density, vapor and temperature fields, while the motion, mass and temperature of the droplets are described using a Lagrangian approach. The computational domain consists of a horizontal cylinder into which the droplet-laden sneezing jet is injected via a circular orifice of radius *R* = 1 *cm* that mimics the mouth opening (52). The cylinder has dimensions *L*_*θ*_ × *L*_*r*_ × *L*_*z*_ = 2*π* × 150*R* × 300*R* = 2*π* × 150 *cm* × 300 *cm* along the azimuthal, radial and axial directions. A total mass of liquid equal to *m*_*l*_ = 8.08 × 10^−6^ *kg* is ejected together with the sneezing jet. The resulting volume fraction is Φ*v* = 4.55 × 10^−6^, in agreement with previous measurements (19, 53). The inflow velocity profile is obtained from a gamma-probability-distribution function (22) and the overall duration of the injection stage is about 0.6 seconds. The jet has a temperature of *T*_*j*_ = 308 *K* and a relative humidity equal to *RH*_*j*_ = 90% (19, 54, 55), while its peak velocity is *u*_*z,j*_ = 20 *m/s* (30, 56). Please note that in the simulation used to compare numerical and experimental results, we consider a jet having the same temperature and humidity of the ambient. For the liquid phase, for each respiratory droplet, its initial diameter is assumed to follow a log-normal distribution with geometric mean equal to 12 *µm* and geometric standard deviation equal to 0.7 (25). The ambient is assumed quiescent and characterized by a uniform temperature and relative humidity and constant thermodynamic pressure. We consider four ambient conditions: two temperatures, *T* = 5 °*C* and *T* = 20 °*C*, and two relative humidities, *RH* = 50% and *RH* = 90%. To simulate the presence of non-volatile elements such as salt, protein and pathogens in the respiratory liquid (57), the minimum size that a droplet can attain has been limited to 3% of the initial volume (≃ 30% of the initial diameter) (24–27).

### Experiments

The experimental setup has been designed to obtain a repeatable droplets-laden jet having properties (jet duration, flow rate) analogous to those considered in the numerical simulations. To prevent exposure of human beings to the potentially harmful laser light, a dummy head is used. The flow is generated by a compressor-based system and is controlled with the aid of an electromagnetic valve. The air stream is seeded with non evaporating, tracer-like droplets (average size 2 *µ*m, Stokes number *St* ≪1) and finally emitted through a circular opening (radius *R* = 1 *cm*) located on the front of the head. The seeding solution is kept at ambient temperature. We observed that the droplets remain suspended in the ambient for long time, without any apparent effect of sedimentation. We performed 94 experiments, consisting of a series of 7 recordings with high speed cameras and 87 velocity measurements with hotwire anemometry. All the experiments are performed in the same flow conditions (fluids temperature, jet duration, flow velocity). We use high-speed imaging system (acquisition rate 0.8 *kHz*) to record the evolution of the flow on a 4 *mm* thick vertical plane. The main components of the imaging system are a double-pulse laser (25 *mJ* per pulse), and a high-speed camera (sensor size of 2560 × 1600 pixel at 0.8 *kHz*) looking perpendicularly to the laser sheet and located at a distance of 2 *m* from the laser plane. The droplet distribution (see inset panels in figure 1) is processed to identify the relevant flow quantities, such as the front and opening angle of the jet. Finally, we employ hot-wire anemometry technique (acquisition rate 1 *kHz*) to characterize the axial flow evolution, i.e. we measure the time- and space-dependent flow axial velocity at different *z* locations. This measurements are also used to verify that the flow generated is highly repeatable.

## Data Availability

Original data created for the study are or will be available in a persistent repository upon publication

## ACKNOWLEDGMENTS

CINECA supercomputing center (Bologna, Italy) is gratefully acknowledged for generous allowance of computer resources under grants HP10C1XSJU. Authors gratefully acknowledge the financial support from the MSCA-ITN-EID project “COMETE” (813948), from PRIN project “Advanced computations and experiments in turbulent multiphase flow” (2017RSH3JY) and from the Research project promoted by China Scholarship Council (201806250023).

