## Supplementary information for "Short-range exposure to airborne virus transmission and current guidelines"

### Your main manuscript title

#### This PDF file includes:

Supplementary text
Figs. S1 to S7 (not allowed for Brief Reports)
Tables S1 to S2 (not allowed for Brief Reports)
Legends for Movies S1 to S4
SI References

#### Other supplementary materials for this manuscript include the following:

Movies S1 to S4

### Supporting Information Text

### Methods: Numerical simulations

Numerical simulations are based on an hybrid Eulerian-Lagrangian framework. An Eulerian approach is used to describe the gaseous phase while a Lagrangian approach is used to track the motion of the respiratory droplets. In the following, the numerical framework, the parameters and the initial and boundary conditions adopted for the simulations will be detailed.

**Description of the gaseous phase.** The velocity, vapor fraction, temperature and density fields of the gaseous phase are described using an Eulerian approach. The governing equations are solved in cylindrical coordinates in an open environment at constant pressure $p_0$. Considering the larger Reynolds number that characterizes a sneezing event (with respect to a cough), a large eddy simulation (LES) approach is employed. Although the choice of a LES approach may reduce the accuracy of the simulations, a posteriori analysis showed that the results obtained are in excellent agreement with those obtained from direct numerical simulations (DNS). Indeed, for the present configuration and considering the grid resolutions employed for the LES, the regions characterized by high values of the viscous dissipation are extremely localized and their contribution to the overall system dynamics is negligible. Under these hypothesis and after applying the Favre-weighted filtering (1) to the asymptotic low-Mach expansion of the Navier-Stokes system, the governing equations read as follows:

$$\frac{\partial \overline{\rho}}{\partial t} + \frac{\partial \overline{\rho}\tilde{u}_i}{\partial x_i} = \overline{S}_m \, , \qquad [1]$$

$$\frac{\partial \overline{\rho}\tilde{u}_i}{\partial t} + \frac{\partial \overline{\rho}\tilde{u}_i\tilde{u}_j}{\partial x_j} = -\frac{\partial \overline{p}}{\partial x_i} + \frac{\partial}{\partial x_j}\left[(\mu_g + \mu_{sgs})\left(\frac{\partial \tilde{u}_i}{\partial x_j} + \frac{\partial \tilde{u}_j}{\partial x_i} - \frac{2}{3}\frac{\partial \tilde{u}_i}{\partial x_i}\delta_{ij}\right)\right] + (\overline{\rho} - \rho_g)g_i + \overline{S}_{p,i} \, , \qquad [2]$$

$$\frac{\partial \overline{\rho}\tilde{Y}_v}{\partial t} + \frac{\partial \overline{\rho}\tilde{Y}_v\tilde{u}_i}{\partial x_i} = \frac{\partial}{\partial x_i}\left(\overline{\rho}(D + D_{sgs})\frac{\partial \tilde{Y}_v}{\partial x_i}\right) + \overline{S}_m \, , \qquad [3]$$

$$\frac{\partial \tilde{u}_i}{\partial x_i} = \frac{\gamma - 1}{\gamma}\frac{1}{p_0}\left[\frac{\partial}{\partial x_i}\left((k_g + k_{sgs})\frac{\partial \tilde{T}}{\partial x_i}\right) + \overline{S}_e - L_v\overline{S}_m\right] \, , \qquad [4]$$

$$\tilde{T} = \frac{p_0}{\overline{\rho}R_g} \, , \qquad [5]$$

where $\overline{\rho}$, $\tilde{u}_i$, $\tilde{Y}_v$, $\tilde{T}$, $\overline{p}$ are the density, velocity, vapor mass fraction, temperature and hydrodynamic pressure fields while $\mu_g$ is the dynamic viscosity of the gaseous phase, $D$ the binary mass diffusion coefficient, $k_g$ the thermal conductivity of the vapor-air mixture and $L_v$ the latent heat of vaporization of the liquid phase. The gaseous phase is assumed to be governed by the equation of state where $R_g = R/W_g$ is the gas constant of the mixture being $W_g$ its molar mass and $R$ the universal gas constant. The ratio $\gamma = c_{p,g}/c_{v,g}$ is the specific heat ratio of the carrier mixture where $c_{p,g}$ and $c_{v,g}$ the gaseous phase specific heat capacity at constant pressure and volume, respectively. In the Navier-Stokes equations, the relative buoyancy force of the jet is accounted via the term $(\overline{\rho} - \rho_g)g_i$ being $\rho_g$ the density of the ambient humid air and $g_i$ the $i$-th component of the gravity acceleration. The subgrid-scale terms of the Navier-Stokes equations are described using the classical Smagorinsky model (2):

$$\mu_{sgs} = \overline{\rho}(C_s\Delta)^2\left\|\frac{1}{2}\left(\frac{\partial \tilde{u}_i}{\partial x_j} + \frac{\partial \tilde{u}_j}{\partial x_i}\right)\right\| \, , \qquad [6]$$

where $C_s$ is a model constant (0.12 in our setup) and $\Delta = [(r\Delta_\theta)\Delta_r\Delta_z]^{1/3}$ is the typical cell size. For the other subgrid-scale fluxes, $D_{sgs}$ and $k_{sgs}$, we adopt the gradient model (3) and their value are assumed proportional to the Smagorinsky eddy-viscosity with a constant turbulent Schmidt and Prandtl numbers equal to $Sc_t = 0.66$ and $Pr_t = 0.78$, respectively.

The effects of the dispersed phase on the gaseous phase are accounted for by three sink-source terms, $\overline{S}_m$, $\overline{S}_{p,i}$ and $\overline{S}_e$:

$$\overline{S}_m = -\sum_{k=1}^{n_d}\frac{dm_k}{dt}\delta(x_i - x_{k,i}) \, , \qquad [7]$$

$$\overline{S}_{p,i} = -\sum_{k=1}^{n_d}\frac{d}{dt}(m_k u_{k,i})\delta(x_i - x_{k,i}) \, , \qquad [8]$$

$$\overline{S}_e = -\sum_{k=1}^{n_d}\frac{d}{dt}(m_k c_l T_k)\delta(x_i - x_{k,i}) \, , \qquad [9]$$

where $x_{k,i}$, $m_k$ and $T_k$ are $k$-th droplet position, mass, velocity and temperature while $c_l$ is the liquid specific heat. The sum is taken over the entire domain droplet population (being $n_d$ the total number of droplets) and, the delta function expresses that the sink-source terms act only at the domain locations occupied by the droplets. These terms are calculated in correspondence of each grid node by volume averaging the mass, momentum, and energy sources from all droplets located within the cell volume centered around the considered grid point.

**Description of the respiratory droplets.** The motion of the respiratory droplets is described using a Lagrangian approach. In particular, considering the small size of the droplets, these are treated as small rigid evaporating spheres and are approximated as point-wise particles. In addition, the temperature of the liquid phase is assumed to be uniform inside each droplet. As the volume (and mass) fraction of the fluid phase in real coughs and sneezes is relatively small (4–6), the mutual interactions among droplets (i.e collisions, coalescence of droplets) can be neglected. Besides, the effect of the subgrid-scale terms is not taken into consideration. Hence, only the resolved part of the Eulerian fields is used in the equations of the dispersed phase. With these assumptions, the position, velocity, mass and temperature of the droplets are described by the following equations:

$$\frac{dx_{k,i}}{dt} = u_{k,i} \, , \qquad [10]$$

$$\frac{du_{k,i}}{dt} = \frac{(\tilde{u}_i - u_{k,i})}{\tau_k}(1 + 0.15Re_k^{0.687}) + (1 - \frac{\overline{\rho}}{\rho_l})g_i \, , \qquad [11]$$

$$\frac{dr_k^2}{dt} = -\frac{\mu_g}{\rho_l}\frac{Sh}{Sc}\ln(1 + B_m) \, , \qquad [12]$$

$$\frac{dT_k}{dt} = \frac{1}{3\tau_k}\left[\frac{Nu}{Pr}\frac{c_{p,g}}{c_l}(\tilde{T} - T_k) - \frac{Sh}{Sc}\frac{L_v}{c_l}\ln(1 + B_m)\right] \, , \qquad [13]$$

where $x_{k,i}$, $u_{k,i}$, $r_k$ and $T_k$ are the position, velocity, radius and temperature of the $k$-th droplet while $\rho_l$ is the liquid droplet density, $c_{p,g}$ the specific heat capacity of the gaseous phase at constant pressure and $L_v$ the latent heat of vaporization. The droplet relaxation time, $\tau_k$, and the droplet Reynolds number, $Re_k$, are defined as:

$$\tau_k = \frac{2\rho_l r_k^2}{9\mu_g} \, , \qquad Re_k = \frac{2\rho_l \|\tilde{u}_i - u_{k,i}\| r_k}{\mu_g} \, , \qquad [14]$$

while the Schmidt number, $Sc$, and Prandtl number, $Pr$, are computed as:

$$Sc = \frac{\mu_g}{\rho_g D} \, , \qquad Pr = \frac{\mu_g c_{p,g}}{k_g} \, , \qquad [15]$$

where $\mu_g$ and $\rho_g$ are the dynamic viscosity and density of the gaseous phase while $D$ is the binary mass diffusion coefficient and $k_g$ the thermal conductivity. The Sherwood number, $Sh$, and Nusselt number, $Nu$, are estimated as a function of the droplet Reynolds number using the Frössling correlations (7):

$$Sh_0 = 2 + 0.552Re_k^{1/2}Sc^{1/3} \, , \qquad Nu_0 = 2 + 0.552Re_k^{1/2}Pr^{1/3} \, . \qquad [16]$$

The resulting Sherwood and Nusselt numbers are corrected to account for the Stefan flow (8, 9):

$$Sh = 2 + \frac{Sh_0 - 2}{F_m} \, , \qquad Nu = 2 + \frac{Nu_0 - 2}{F_t} \, . \qquad [17]$$

The coefficients $F_m$ and $F_t$ are computed as follows:

$$F_m = \frac{(1 + B_m)^{0.7}}{B_m}H_m \, , \qquad F_t = \frac{(1 + B_t)^{0.7}}{B_t}H_t, \qquad [18]$$

where $H_m$ and $H_t$ are defined as:

$$H_m = \ln(1 + B_m) \, , \qquad H_t = \ln(1 + B_t) \, , \qquad [19]$$

being $B_m$ and $B_t$ the Spalding mass and heat transfer numbers:

$$B_m = \frac{Y_{v,s} - \tilde{Y}_v}{1 - Y_{v,s}} \, , \qquad B_t = \frac{c_{p,v}}{L_v}(\tilde{T} - T_k) \, , \qquad [20]$$

where $\tilde{Y}_v$ and $\tilde{T}$ are the vapor mass fraction and temperature fields evaluated at the droplet position, $Y_{v,s}$ is the vapor mass fraction evaluated at droplet surface and $c_{p,v}$ is the vapor specific heat at constant pressure. The vapor mass fraction at the droplet surface corresponds to the mass fraction of the vapor in a saturated vapor-gas mixture at the droplet temperature. To estimate $Y_{v,s}$, we use the Clausius-Clapeyron relation to first compute the vapor molar fraction, $\mathcal{X}_{v,s}$:

$$\mathcal{X}_{v,s} = \frac{p_{ref}}{p_0}\exp\left[\frac{L_v}{R_v}\left(\frac{1}{T_{ref}} - \frac{1}{T_k}\right)\right] \, , \qquad [21]$$

where $p_{ref}$ and $T_{ref}$ are arbitrary reference pressure and temperature and $R_v = R/W_l$ is the vapor gas constant. The saturated vapor mass fraction is then computed using the relation:

$$Y_{v,s} = \frac{\mathcal{X}_{v,s}}{\mathcal{X}_{v,s} + (1 - \mathcal{X}_{v,s})\frac{W_g}{W_l}} \, , \qquad [22]$$

where $W_g$ and $W_l$ are are the molar mass of the gaseous and liquid phases.

**Numerical method.** The numerical code consists of two different modules: i) an Eulerian module that solves the governing equations for the gaseous phase (density, velocity, vapor mass fraction and temperature); ii) a Lagrangian module that solves the equations governing the droplet dynamics (position, velocity, mass and temperature). In particular, the governing equations of the gaseous phase are discretized in space using a second-order central finite differences scheme and they are time advanced using a low-storage third-order Runge-Kutta scheme. Likewise, the governing equations of the droplets are time integrated using the same Runge-Kutta scheme, and a second-order accurate polynomial interpolation is used to evaluate the Eulerian quantities at the droplet position. Please refer to previous works (9–11) for additional validations and tests of the numerical method.

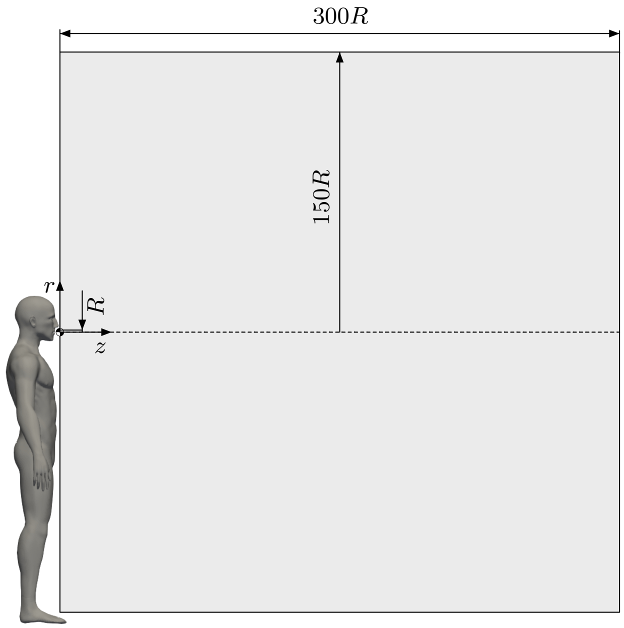


**Fig. S1.** Sketch of the simulation setup used for the simulations. The computational domain is a cylinder having dimensions $L_\theta \times L_r \times L_z = 2\pi \times 150R \times 300R$ being $R = 1\ cm$ the radius of the circular orifice that mimics the mouth opening. The sneezing jet, together with the respiratory droplets, are injected from the left side of the domain (through the orifice). The domain is initially quiescent (zero velocity) and characterized by a uniform value of temperature, humidity.

**Simulation setup.** The computational domain, figure S1, is a cylinder into which the droplet-laden sneezing jet is injected through a circular orifice of radius $R = 1\ cm$ located at the centre of the left base that mimics the average mouth opening for females and males subjects (4, 12). The cylinder dimensions are $L_\theta \times L_r \times L_z = 2\pi \times 150R \times 300R$ along the azimuthal ($\theta$) radial ($r$) and axial ($z$) directions. The domain is discretized using a staggered grid with $N_\theta \times N_r \times N_z = 96 \times 223 \times 1024$ grid points.

A total mass of liquid equal to $m_l = 8.08 \times 10^{-6}\ kg$ is ejected together with the sneezing jet; the mass of the ejected gaseous phase is equal to $m_g = 2.00 \times 10^{-3}\ kg$. The resulting mass fraction is equal to $\Phi_m = 4.04 \times 10^{-3}$ while the volume fraction is equal to $\Phi_v = 4.55 \times 10^{-6}$ conforming to previous experimental studies (4–6, 13, 14).

The inflow velocity profile of the sneezing jet (figure S2) is obtained from a gamma-probability-distribution function (15) and a simple conversion from dynamic pressure to velocity is implemented based on Bernoulli's principle. The overall duration of the injection stage (sneezing jet and droplets) is about 0.6 $s$ (15). The sneezing jet is characterized by a temperature of $T_j = 308\ K$ and a relative humidity $RH_j = 90\%$ (4, 16, 17) and its peak velocity is $u_{z,j} = 20\ m/s$ (18, 19). From the temperature and relative humidity of the sneezing jet, the density and vapor mass fraction of the jet are obtained from the revised formula reported in Picard *et al.* (2008)(20). The other thermo-physical and transport properties are estimated from Tsilingiris (2008) (21), see table S2 for details.

Considering now the liquid phase (respiratory droplets), for each droplet injected in the computational domain, its initial diameter is assumed to follow a log-normal distribution with geometric mean equal to 12 $\mu m$ and geometric standard deviation ($GSD$) equal to 0.7 (22). The above mentioned distribution is generated using a Gaussian random number generator based on a Ziggurat method (23). The initial velocity of the droplets is obtained through interpolation of the velocity field of the gaseous phase in the inlet region, while the initial temperature of the droplets is set to $T = 308\ K$. To mimic the presence of salt, protein and virus dissolved in the respiratory droplets (24), the droplets have a non-volatile core and thus they cannot completely evaporate. This leads to the formation of the so-called droplet nuclei, i.e. the residual part of the respiratory droplets that does not evaporate. In agreement with previous studies (22, 25–29), we consider that the non-volatile core of each droplet represents the 3% of the droplet volume. In terms of droplets size, this means that a droplet can shrink down to $\simeq 30\%$ of its initial diameter. Due to numerical stability issues (9), all generated droplets with an initial size smaller than a critical radius of 0.65 $\mu m$ will be treated as tracers. Likewise, if a droplet, due to the evaporation, becomes smaller than the critical radius, it will be treated as a tracer.

The ambient is assumed quiescent (i.e. all velocity components are initially set to zero) and is characterized by a uniform value of temperature, humidity and constant thermodynamic pressure. The density of the gaseous phase is obtained from the gas equation of state, while the vapor mass fraction is obtained through the Clausius–Clapeyron relation using the sneezing jet conditions as reference.

We perform a total of 5 simulations: a benchmark simulation used for the comparison with the experiments (case S0 in table S2) and four production simulations (cases S1-4 in table S2). The benchmark case considers mono-dispersed non-evaporating droplets (diameter of 2 $\mu m$) released in a sneezing jet having the same temperature of the ambient: $T = 295\ K$ (22 $°C$) and humidity: $RH = 50\%$ (TU Wien laboratory conditions). The other four simulations consider different ambient conditions: two temperatures, $T = 278\ K$ (5 $°C$) and $T = 293\ K$ (20 $°C$), and two relative humidity values, $RH = 50\%$ and $RH = 90\%$. A detailed summary of the simulation parameters and thermo-physical properties adopted for the different simulations is reported in table S1-S2.

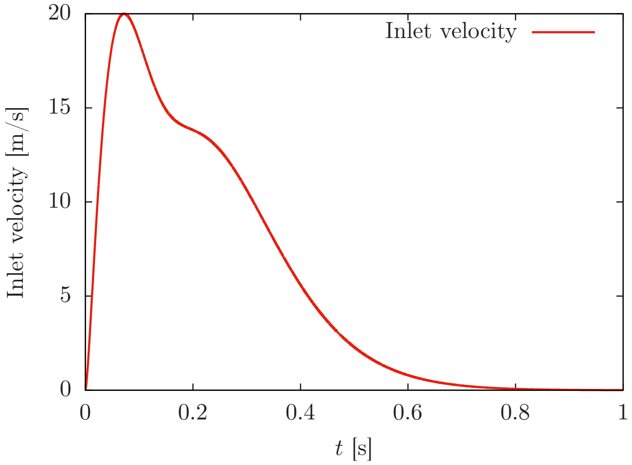


**Fig. S2.** Inflow velocity of the sneezing jet used in the simulations. The inlet velocity is obtained from a gamma-probability-distribution function (15). The duration of the sneezing event is $\simeq 0.6\ s$ and the peak velocity is $20\ m/s$.

**Table S1. Summary of the simulation parameters and thermophysical properties.**

| Parameter | Symbol | Value | Unit of measurement |
|---|---|---|---|
| Inlet radius | R | 1.00 $\times$ 10$^{-2}$ | m |
| Sneezing jet temperature | $T_j$ | 308 | K |
| Sneezing jet relative humidity | $RH_j$ | 90% | - |
| Maximum sneezing jet velocity | $u_{z,j}$ | 20 | m/s |
| Droplets temperature | $T_k$ | 308 | K |
| Mass injected liquid phase | $m_l$ | 8.08 $\times$ 10$^{-6}$ | kg |
| Mass injected gaseous phase | $m_g$ | 2.00 $\times$ 10$^{-3}$ | kg |
| Liquid mass fraction | $\Phi_m$ | 4.04 $\times$ 10$^{-3}$ | - |
| Liquid volume fraction | $\Phi_v$ | 4.55 $\times$ 10$^{-6}$ | - |
| Environment temperature | $T$ | 278 and 293 | K |
| Environment relative humidity | $RH$ | 50% and 90% | - |
| Environment thermodynamic pressure | $p_0$ | 1.01 $\times$ 10$^{5}$ | Pa |
| Dynamic viscosity gaseous phase | $\mu_g$ | 1.99 $\times$ 10$^{-5}$ | Pa s |
| Thermal conductivity gaseous phase | $k_g$ | 2.63 $\times$ 10$^{-2}$ | W/(m*K) |
| Latent heat of vaporization | $L_v$ | 2.41 $\times$ 10$^{6}$ | J/kg |
| Universal gas constant | $R$ | 2.87$\times$ 10$^{2}$ | J/(kg*K) |
| Molar mass of the gaseous phase | $W_g$ | 2.89 $\times$ 10$^{-2}$ | kg/mol |
| Gas constant gaseous phase | $R_g$ | 2.92 $\times$ 10$^{2}$ | J/(kg*K) |
| Specific heat capacity at constant pressure gaseous phase | $c_{p,g}$ | 1.03 $\times$ 10$^{3}$ | J/(kg*K) |
| Specific heat capacity at constant volume gaseous phase | $c_{v,g}$ | 7.42 $\times$ 10$^{2}$ | J/(kg*K) |
| Specific heat ratio gaseous phase | $\gamma$ | 1.39 | - |
| Vapor specific heat capacity at constant pressure | $c_{p,v}$ | 1.88 $\times$ 10$^{3}$ | J/(kg*K) |
| Vapor phase gas constant | $R_v$ | 4.61 $\times$ 10$^{2}$ | J/(kg*K) |
| Binary mass diffusion coefficient | D | 2.67 $\times$ 10$^{-5}$ | m$^2$/s |
| Molar mass liquid phase | $W_l$ | 1.80 $\times$10$^{-2}$ | kg/mol |
| Density liquid phase | $\rho_l$ | 1.00 $\times$ 10$^{3}$ | kg/m$^3$ |
| Specific heat liquid phase | $c_l$ | 4.18 $\times$ 10$^{3}$ | J/(kg*K) |
| Volume fraction non-volatile material droplet | $\Phi_v^c$ | 3% | - |
| Prandtl number | $Pr$ | 0.782 | - |
| Schmidt number | $Sc$ | 0.663 | - |

**Table S2. Thermophysical properties of the gaseous phase (ambient)**

| Case | Temperature $T$ [K] | Relative humidity $RH$ [%] | Density $\rho_g$ [kg/m$^3$] | Vapor mass fraction $Y_g$ |
|---|---|---|---|---|
| S0 | 295 | 50 | 1.174 | 8.22 $\times 10^{-3}$ |
| S1 | 278 | 50 | 1.245 | 2.82 $\times 10^{-3}$ |
| S2 | 278 | 90 | 1.245 | 5.09 $\times 10^{-3}$ |
| S3 | 293 | 50 | 1.181 | 7.42 $\times 10^{-3}$ |
| S4 | 293 | 90 | 1.181 | 1.34 $\times 10^{-2}$ |

### Methods: Experiments

We set up a laboratory experiment to investigate the dynamics of droplets-laden jets. We used a compressor-based system to supply the flow with air, which is seeded with micrometrical droplets by a liquid seeder. Measurements consist of flow velocity (point wise) and drops distribution (two-dimensional distribution). Details are provided in the following.

**Experimental setup.** The main components of the system are shown in figure S3(a). To produce repeatable flow conditions, we designed a system in which the flow parameters (pressure, duration) can be carefully controlled. The flow generated by the compressor (pressure 6.5 bar) is controlled by an electromagnetic valve (Parker 4818653D D5L F). The valve is activated by a timer (Finder, relays type 94.02 and plug-in timer 85.02), which is set to maintain the valve open for 0.15 $s$. We verified *a-posteriori* via hot-wire measurements that the flow is highly repeatable.

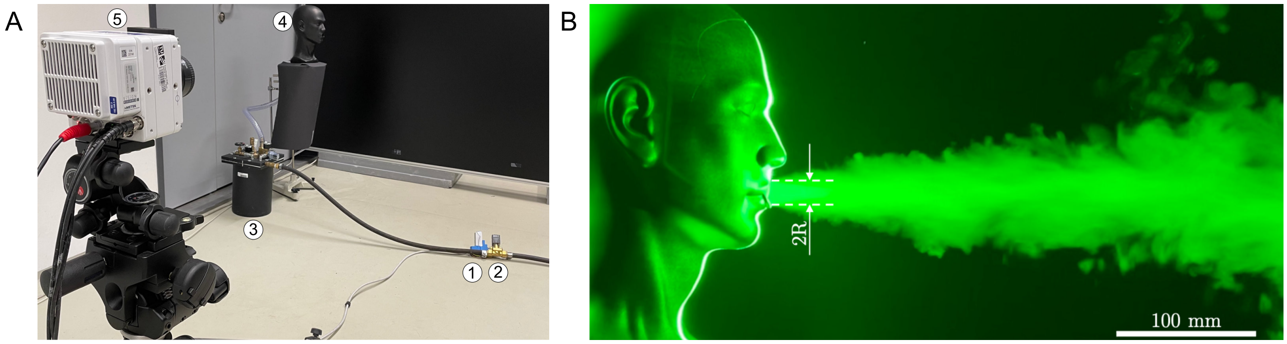


**Fig. S3.** Panel A shows the experimental setup used. The setup is composed by the compressor (not shown), timer (1), electromagnetic valve (2), liquid seeder (3) and dummy head (4). A laser is used to illuminate the micro-metric droplets. Image acquisition is preformed by a high-speed camera (5). Panel B shows the dummy head used to perform the experiments ($R = 1\ cm$).

The compressor is connected to a seeding generator (9010F0031 Liquid Seeder, type FT700CE), which produces droplet with size falling in the range 1-3 $\mu$m, with an average droplets size of 2 $\mu$m, as reported in figure S4. To seed the flow with neutrally-buoyant and non-evaporating drops, an aqueous and non-toxic solution (Safex - Inside Nebelfluid, Dantec Dynamics) is used. The solution is kept at the ambient temperature. We observed that the drops remain suspended in the ambient for long time, without any apparent effect of sedimentation. The droplets Stokes number, $St$, is defined as $St = \rho_l r_l^2 \bar{u}_{z,j}/(R\mu_g)$, being $r_l = 1\ \mu$m averaged droplets radius and $\bar{u}_{z,j} \leq 20$ m/s the reference velocity. For the present case, we obtain $St \leq 0.1$ and we consider the droplets as flow tracers (30).

We used a dummy head to avoid exposure of human beings to the potentially harmful laser light. The droplet-laden jet is emitted through the mouth of the dummy, which is mimicked by a circular opening of radius $R = 1$ cm, see figure S3(B). The mouth is directly connected to the fog generator through a tube of length 100 $cm$ and inner diameter $2R$. The temperature of environment ($T$), jet ($T_j$) and droplets ($T_k$) is constant and equal to $T = T_j = T_k = 295$ K, therefore buoyancy plays no role in the dynamics of the jet, in agreement with the observations of (31). The relative humidity of the air is $RH = 50\%$.

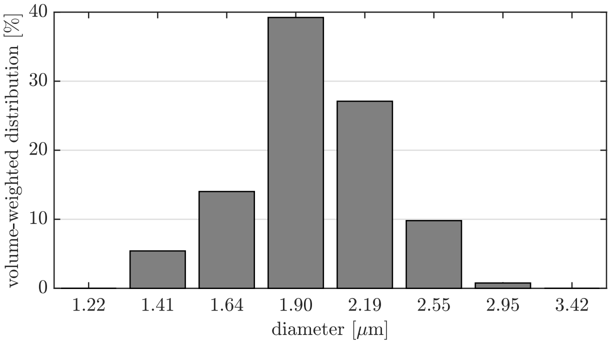


**Fig. S4.** Volume-weighted droplets distribution [%] as a function of the diameter of the droplets produced (32).

**Imaging system.** A high-speed laser is used to create a sheet (thickness 4 $mm$) in which the experimental measurements are performed. The laser consists of a double-pulse laser (Litron LD60-532 PIV, 25 m$J$ per pulse) illuminating the measurement region at frequency 0.8 $kHz$. To record the evolution of the flow, we used a Phantom VEO 340L (sensor size of $2560 \times 1600$ pixel at 0.8 kHz) equipped with lenses having focal length 35 $mm$, looking perpendicularly to the laser sheet at a distance of 200 $cm$. Camera and laser are controlled via a high-speed synchroniser (PTU X, LaVision GmbH, Germany). Images are collected with DAVIS 10 (LaVision GmbH, Germany) and processed in MATLAB to compute the extension of the front of the jet.

As the jet propagates along the axial direction ($z$), particles concentration reduces. As a result, the light intensity recorded by the cameras drops significantly with $z$, making the detection of the front of the jet hard to obtain. To perform the edge-detection process, we applied subsequent image processing steps (subtracting background noise, binarization, median filter). After image preprocessing, the boundary of the jet is found by Moore-Neighbor tracing algorithm (33) and finally the edge, the maximum horizontal coordinate of the boundary, is determined and tracked in time. In figure S5 (and as well in the manuscript), the evolution of the front of the jet, $L$, is reported as a function of time. The mean (red solid line) and standard deviation (error bars) are obtained from 7 independent experiments.

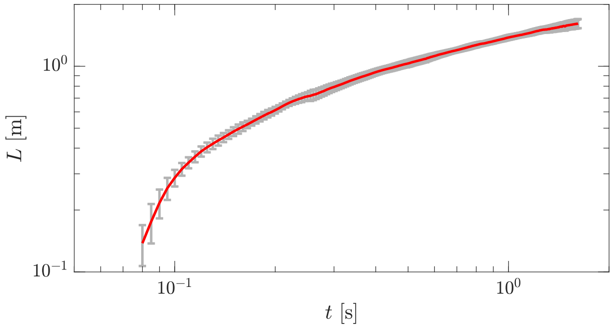


**Fig. S5.** Evolution of the front of the jet, $L$, as a function of time, $t$. Mean (red solid line) and standard deviation (error bars) are obtained from 7 independent experiments. The acquisition rate is 0.8 kHz and we show here one every 4 instants.

**Hot-wire anemometry.** We used a hot-wire anemometry system (acquisition rate 1 $kHz$, probe type Dantec 55P11) to measure the axial velocity of the flow and thus to calibrate the inflow velocity profile. Figure S6 shows the axial velocity measured along the centerline of the jet ($r = 0$) at a distance $z = 20$ mm from the mouth as a function of time, $t$. To characterise the inlet condition, we performed 11 independent experiments. In each experiment, the instantaneous velocity measurements ($u_1$, grey data) are averaged over a moving window of 20 ms to obtain $u_{20}$ (blue solid line). Then results of all experiments are averaged to obtained the ensemble averaged flow velocity ($u$, red solid line). The excellent agreement observed between the ensemble average ($u$) and the single experiment ($u_{20}$) confirms the repeatability of the flow generated. Finally, the mean value of velocity computed for $0 \leq t \leq 0.7$ $s$ ($\overline{u}$, dashed line) is also shown, and it is used for further comparison with the results obtained from the numerical simulations.

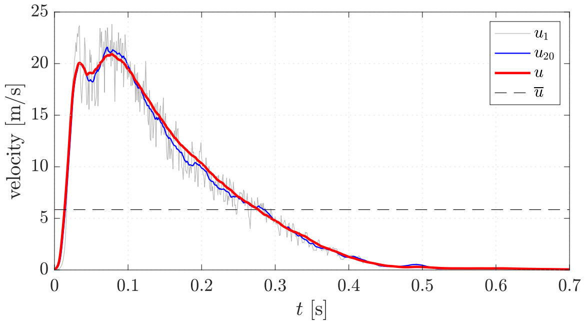


**Fig. S6.** Time-dependent evolution of the axial velocity measured at the centerline ($r = 0$) at distance $z = 20$ mm from the mouth. Instantaneous velocity measurements ($u_1$, grey line) as well as the velocity averaged over a moving window of 20 ms ($u_{20}$, blue line) are shown here for one experiment. Then results of 11 experiments are used to obtained the ensemble averaged flow velocity ($u$, red solid line). Finally, the mean value computed for $0 \leq t \leq 0.7$ $s$ is shown ($\overline{u}$, dashed line).

### Comparison between simulations and experiments

In addition to the comparison between experiments and simulations reported in the manuscript (front of the jet and semi-cone angle, see figure 1), to further benchmark experimental results against numerical results, we investigate the evolution of the time-averaged jet velocity, $\overline{u}$, along the axial direction ($z$ axis). The time interval used for the average spans from $t = 0$ $s$ up to $t = 0.7$ $s$. Concerning the experiments, 76 independent realizations are used to determine $\overline{u}(z)$, which is defined as described above. Measurements are performed along the centerline at 76 equally-spaced $z$ positions. For simulations, data are obtained from the Eulerian grid used to compute the velocity fields and then averaged in time. Results are shown in figure S7 for simulations (red dots) and experiments (blue dots). The velocity profiles are reported normalized by $\overline{u}_0$, i.e. the first velocity value (closest point to the inlet position). The scaling law, $\overline{u} \propto 1/x$, is also reported as a reference (34) with a black, dashed line. As can be appreciated from the figure, experiments and simulations are in excellent agreement over the entire axis span. In addition, both experimental and numerical results well match with the analytic scaling law.

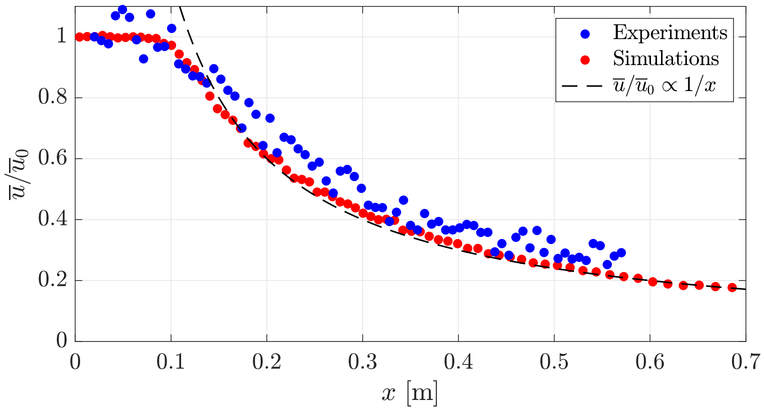


**Fig. S7.** Time-averaged axial velocity, $\overline{u}$, measured at the jet centerline and normalised by the velocity at the closet point to the inlet position ($\overline{u}_0$), is reported as a function of the distance from the mouth, $z$. Experimental results are represented using blue dots while simulations results with red dots. As a reference, the scaling law $\overline{u}/\overline{u}_0 \propto 1/x$ (dashed, black line) is also reported.

**Movie S1. Movie showing the first 3 seconds of a sneezing event for $T = 5$ °C and $RH = 50\%$. The background shows the local value of the relative humidity (black-low; white-high). The respiratory droplets are displayed rescaled according to their diameter (not in real scale) and are also colored according to their size (red-small; white-large). From the movie, the presence of localized supersaturated regions where $RH > 100\%$ (white) can be appreciated. The upward motion of the sneeze cloud produced by buoyancy and, of part of the respiratory droplets, can be also appreciated.**

**Movie S2. Movie showing the first 3 seconds of a sneezing event for $T = 5$ °C and $RH = 90\%$. The background shows the local value of the relative humidity (black-low; white-high). The respiratory droplets are displayed rescaled according to their diameter (not in real scale) and are also colored according to their size (red-small; white-large). For this setting (low temperature and high relative humidity), large supersaturated regions ($RH > 100\%$) can be observed. Droplets present in these regions, due to the local humidity conditions, can possibly grow in size (condensation) instead of shrinking.**

**Movie S3. Movie showing the first 3 seconds of a sneezing event for $T = 20$ °C and $RH = 50\%$. The background shows the local value of the relative humidity (black-low; white-high). The respiratory droplets are displayed rescaled according to their diameter (not in real scale) and are also colored according to their size (red-small; white-large). For this configuration (moderate temperature and low humidity), supersaturated regions are only observed in the beginning of the sneezing event. Nevertheless, most of the droplets are located in regions with a local relative humidity value larger than the ambient. Thus their evaporation dynamics is much slower than predicted by analytic models (e.g. $d^2$-law).**

**Movie S4. Movie showing the first 3 seconds of a sneezing event for $T = 20$ °C and $RH = 90\%$. The background shows the local value of the relative humidity (black-low; white-high). The respiratory droplets are displayed rescaled according to their diameter (not in real scale) and are also colored according to their size (red-small; white-large). Also for this setting, the larger temperature (with respect to $T = 5$ °C) limits the extension of the supersaturated regions even though the ambient humidity is close to the saturation value.**
